# Change in COVID-19 risk over time following vaccination with CoronaVac: A test-negative case-control study

**DOI:** 10.1101/2021.12.23.21268335

**Authors:** Matt D.T. Hitchings, Otavio T. Ranzani, Margaret L. Lind, Murilo Dorion, Tatiana Lang D’Agostini, Regiane Cardoso de Paula, Olivia Ferreira Pereira de Paula, Edlaine Faria de Moura Villela, Mario Sergio Scaramuzzini Torres, Silvano Barbosa de Oliveira, Wade Schulz, Maria Almiron, Rodrigo Said, Roberto Dias de Oliveira, Patricia Vieira da Silva, Wildo Navegantes de Araújo, Jean Carlo Gorinchteyn, Natalie E. Dean, Jason R. Andrews, Derek A.T. Cummings, Albert I. Ko, Julio Croda

## Abstract

**Objective:** To estimate the change in odds of covid-19 over time following primary series completion of the inactivated whole virus vaccine, CoronaVac (Sinovac Biotech) in São Paulo state, Brazil.

**Design:** Test negative case-control study.

**Setting:** Community testing for covid-19 in São Paulo state, Brazil.

**Participants:** Adults aged 18-120 years who were residents of São Paulo state, without a previous laboratory-confirmed covid-19 infection, who received only two doses of CoronaVac, and underwent reverse transcription polymerase chain reaction (RT-PCR) testing for SARS-CoV-2 from 17 January to 30 September 2021.

**Main outcome measures:** RT-PCR-confirmed symptomatic covid-19 and associated hospital admissions and deaths. Cases were pair-matched to test-negative controls by age (in 5-year bands), municipality of residence, healthcare worker (HCW) status, and date of RT-PCR test (±3 days). Conditional logistic regression was adjusted for sex, number of covid-19-associated comorbidities, race, and previous acute respiratory infection.

**Results:** From 137,820 eligible individuals, 37,929 cases with symptomatic covid-19 and 25,756 test-negative controls with covid-19 symptoms were formed into 37,929 matched pairs. Adjusted odds ratios of symptomatic covid-19 increased with time since series completion, and this increase was greater in younger individuals, and among HCWs compared to non-HCWs. Adjusted odds ratios of covid-19 hospitalisation or death were significantly increased from 98 days since series completion, compared to individuals vaccinated 14-41 days previously: 1.40 (95% confidence interval 1.09 to 1.79) from 98-125 days, 1.55 (1.16 to 2.07) from 126-153 days, 1.56 (1.12 to 2.18) from 154-181 days, and 2.12 (1.39-3.22) from 182 days.

**Conclusions:** In the general population of São Paulo state, Brazil, an increase in odds of moderate and severe covid-19 outcomes was observed over time following primary series completion with CoronaVac.

**What is already known on this topic:**

- The effectiveness of the inactivated whole virus vaccine, CoronaVac (Sinovac Biotech) against moderate and severe covid-19 has been demonstrated in clinical trials and observational studies.
- Observational studies have suggested that effectiveness of other covid-19 vaccines appears to decrease over time, prompting many countries to deploy additional doses for individuals who have completed their primary series.
- There is currently no evidence for change in the rate of breakthrough infection in individuals who have received a primary series of CoronaVac.

**What this study adds:**

- In individuals receiving two doses of CoronaVac, the odds of symptomatic covid-19 increased over time since series completion.
- Larger increases in covid-19 odds were observed in individuals aged 18-40, and in healthcare workers compared to non-healthcare workers.
- Odds of covid-19 hospitalisation or death increased over time since series completion, but to a lesser extent.

## Introduction

Since the authorisation and licensing of the first COVID-19 vaccines in late 2020, more than 8 billion doses have been distributed worldwide(1), of which nearly 2 billion have been CoronaVac, an inactivated vaccine produced by the Chinese pharmaceutical Sinovac(2). A key driver of future dynamics of COVID-19, as well as public health policy, will be the durability of vaccine protection against infection and severe disease.

Following large-scale vaccination campaigns, some observational studies have found evidence for waning of effectiveness of COVID-19 vaccines against infection, symptomatic COVID-19, and severe outcomes(3–10). However, there is a lack of data concerning waning from inactivated vaccines, such as CoronaVac, which is urgently needed to guide public health policy. Together with its widespread use, inactivated vaccines elicit lower neutralising antibody response than other vaccine platforms in the short- and long-term period after two doses, reaching almost undetectable levels after 6 months(11). Observational studies of waning effectiveness can be afflicted by multiple sources of bias, including differential build-up of immunity among unvaccinated individuals(12), and association between time of vaccination and COVID-19 risk in risk-prioritised vaccination campaigns. These biases could result in overestimation of waning. In this study, we utilise a well-characterised, linked surveillance database in São Paulo State, Brazil to conduct a matched test-negative case-control study among vaccinated individuals, with the aim of estimating a change in vaccine effectiveness over time following completion of the two-dose schedule of CoronaVac.

## Methods

The study was approved by the Ethical Committee for Research of Federal University of Mato Grosso do Sul (CAAE: 43289221.5.0000.0021). The Free and Informed Consent form was waived because the study involved de-identified surveillance datasets.

### Study setting

The study setting and design have been described in detail elsewhere(13–15). Individual-level data on demographic and clinical characteristics, SARS-CoV-2 testing, and COVID-19 vaccination were assembled from four databases: the São Paulo State laboratory testing registry (GAL), national surveillance databases covering acute respiratory illness (ARI) (e-SUS) and severe ARI (SIVEP-Gripe), and the São Paulo State vaccine registry (Vacina Já) covering all individuals vaccinated in São Paulo State. The surveillance databases include hospitalisations and all other health visits conducted through public and private health systems, and notification of SARS-CoV-2 test results and suspected COVID-19 cases, hospitalisations, and deaths is compulsory. We retrieved information from these databases on October 15^th^, 2021. The STROBE checklist is shown in Supplementary Table 1.

### Study population and design

The study population consisted of adults ≥18 years old who had a residential address in São Paulo State and had complete and consistent information between data sources on age, sex, residence, and vaccination and testing status and dates. Eligible cases had a symptomatic illness, received a positive SARS-CoV-2 RT-PCR test during the study period of 17 January 2021 to 30 September 2021 with sample collection date within 10 days after symptom onset, were without a prior positive SARS-CoV-2 RT-PCR test since January 2020, and received two doses of CoronaVac prior to sample collection. Eligible controls had a symptomatic ARI, received a negative SARS-CoV-2 RT-PCR test during the study period with sample collection date within 10 days after symptom onset, were without a prior positive SARS-CoV-2 RT-PCR test since January 2020, or in the following 14 days, and received two doses of CoronaVac prior to sample collection. Each individual could contribute up to four different negative tests as controls. To control for predictors of the timing of vaccination and changes in infection risk and variant circulation over time and space, we matched each case to a single control by date of RT-PCR testing (±3 days), age (in 5-year bands), municipality of residence (n=645 municipalities), and vaccination priority group for any dose (categorised as healthcare worker vs. not healthcare worker, which included elderly, teachers, general population, etc.). Each control RT-PCR test could serve as a control for multiple cases, and individuals that contributed case events with negative tests prior to their positive test were eligible to serve as a control.

### Outcomes and covariates

We estimated the association between time from receipt of second CoronaVac vaccine dose to RT-PCR sample collection date with the odds of testing positive for SARS-CoV-2. Days between second dose of vaccination and RT-PCR sample collection were categorised in the following time intervals: 0-13, 14-41 (selected as the reference group, when IgG seropositivity peaks(16)), 42-69, 70-97, 98-125, 126-153, 154-181, and ≥182 days. In this design, an odds ratio greater than one can be interpreted as evidence of lower effectiveness than during the reference period of 14-41 days(12,17).

In addition, we estimated the association between days from second dose of vaccination until RT-PCR sample collection and the odds of COVID-19 hospitalisation or COVID-19 death, as a composite outcome. To perform this analysis, we selected only matched pairs in which the case had the outcome of interest and fit the primary model to this subset(14). In this way, controls represented negative RT-PCR tests from ambulatory and hospital settings, with a range of severity, but representative of the population with access to RT-PCR testing(18). We performed analyses within subgroups defined by age (18-39 years, 40-64 years, 65-79 years, and ≥80 years) and by priority status (healthcare worker vs. non-healthcare worker).

We accounted for the following covariates as potential confounders: age as a linear term, sex, self-reported race (brown, black, yellow, white, or indigenous(19)), prior ARI as a measure of healthcare-seeking behaviour (defined as at least one previous symptomatic event that was reported to surveillance systems between 1 February 2020 and 16 January 2021), number of COVID-19-associated comorbidities documented at the time of the RT-PCR test (cardiovascular, renal, neurological, haematological, or hepatic comorbidities, diabetes, chronic respiratory disorder, obesity, or immunosuppression; categorised as none, one-two, and three or more).

### Statistical analysis

We performed conditional logistic regression to estimate the association between days since vaccination and each outcome, accounting for the matched design and including potential confounders as additional covariates. We included a missing indicator for self-reported race. As there was a strong association between age and HCW status (the vast majority of HCWs were under 65), we fit three models with interactions (age by time since vaccination, HCW status by time since vaccination, and both interactions) and used AIC to select the best-fitting model.

### Sensitivity analyses

The test-negative case-control design can exhibit several biases which could lead to spurious conclusions of waning effectiveness. Therefore, we performed a number of sensitivity analyses to detect bias.

1. Individuals vaccinated earlier are likely at higher risk of SARS-CoV-2 infection or developing COVID-19, hence the adjustment for age and prioritisation group. Even so, differences may persist in the risk of RT-PCR-confirmed COVID-19 for those vaccinated earlier versus later. This would lead to apparent waning, as “early adopters”, vaccinated early due to their perceived or occupational risk of exposure to SARS-CoV-2, would be over-represented in later time periods since vaccination and be at increased risk of SARS-CoV-2. To minimise the potential for waning while focusing on differences in risk by calendar time of vaccination, we conducted a sensitivity analysis restricting the study population to individuals who received an RT-PCR test within 14-90 days of receipt of their second dose, a period in which less waning is expected. We matched on time of testing as above, and estimated the odds ratio of testing positive by month of vaccine series completion, relative to April, the month in which the majority of 65-79 year olds were vaccinated. If changes in effectiveness are due to differences in COVID-19 risk by month of vaccination, we would expect to observe elevated odds ratios in earlier months relative to later months. On the other hand, if changes in effectiveness were due to waning, which we expect to be more limited before 90 days, we should observe little difference in COVID-19 odds by month of vaccination. Analyses were performed by age and healthcare worker subgroup.
2. Individuals who are vaccinated later are more likely to have been infected prior to vaccination, due to a longer period of exposure prior to vaccine-conferred protection. Individuals vaccinated later may therefore be less susceptible to infection, which could induce apparent waning by cohort effects when evaluating risk by time since vaccination. To reduce this depletion of susceptibles during the time period of vaccination among HCWs, and restrict to “early adopter” HCWs, we performed the primary analysis on a population with an additional eligibility criteria that they be healthcare workers who received the second dose in February 2021.
3. Finally, if an increased odds of breakthrough infection observed in the primary analysis were due to waning effectiveness alone, we would expect the waning pattern to be the same among those completing their series in February and March. Therefore, among HCWs we compared the pattern of waning among individuals who received the second dose in February 2021 (the “early adopter” HCWs in sensitivity analysis 2) with the pattern among individuals who received the second dose in March 2021.

## Results

### Study setting

The second COVID-19 wave in São Paulo State peaked in March 2021, with substantial declines in transmission from June 2021. This decline is reflected in the decreased proportion of RT-PCR tests that are positive over time in the study population (Supplementary Figure 1).

### Study population

Among 137,840 individuals with 138,720 RT-PCR tests eligible for selection as a case or control, 75,858 RT-PCR tests from 63,651 individuals were matched into 37,929 case-control pairs for the primary analysis. Figure 1 shows the flowchart of enrollment, while Figure 2 shows the timing of vaccination by age group and healthcare worker status. Table 1 shows demographic and clinical characteristics for matched cases and controls. There was a strong correlation between age and HCW priority status; over 98% of HCWs were under 65. The majority of discordant case-control pairs were vaccinated close in time (Supplementary Table 2).

**Figure 1.**
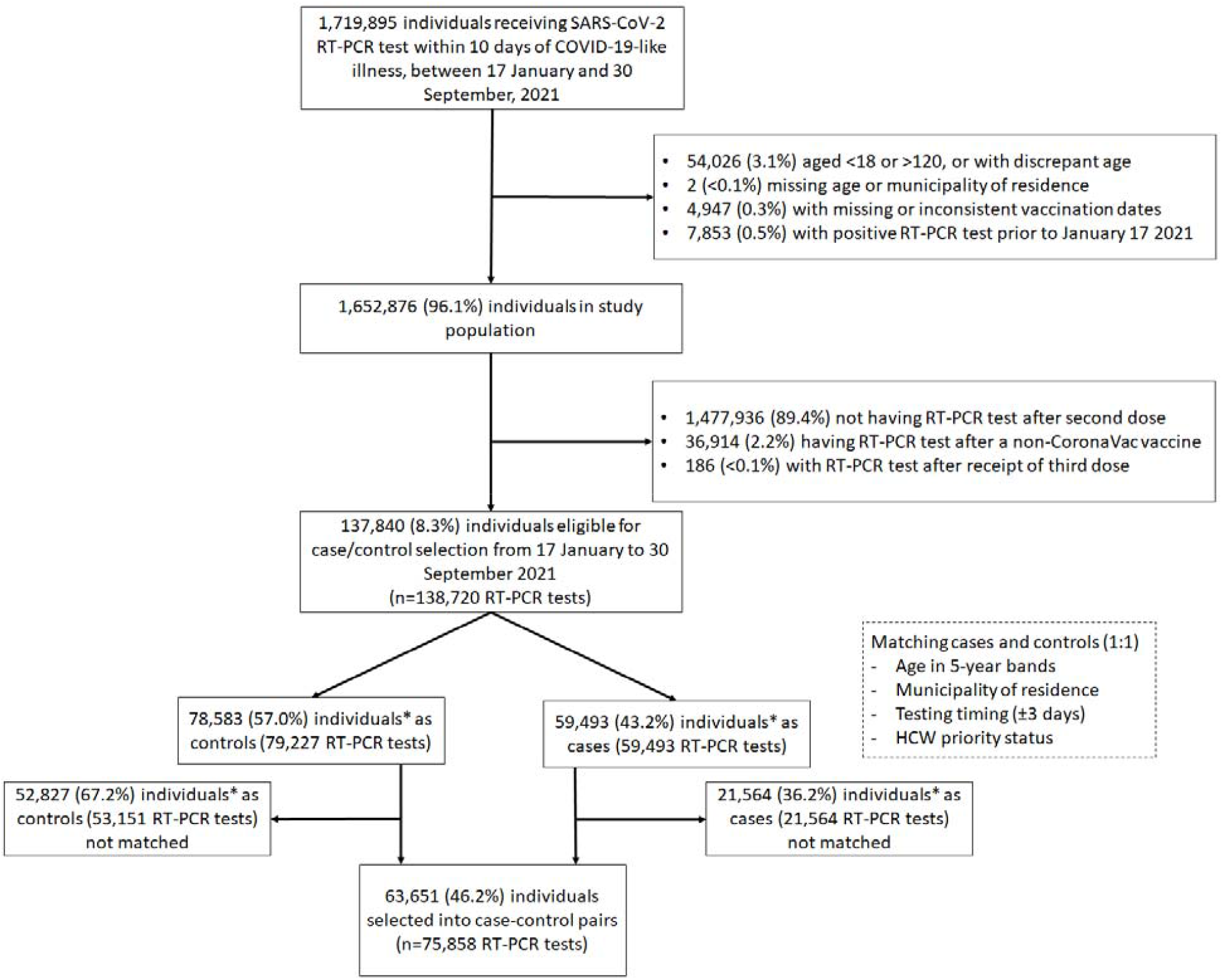
Flow chart of study population and case and control selection *34 participants contributed as controls and cases, and 3,615 RT-PCR tests were used as controls for multiple cases.

**Figure 2.**
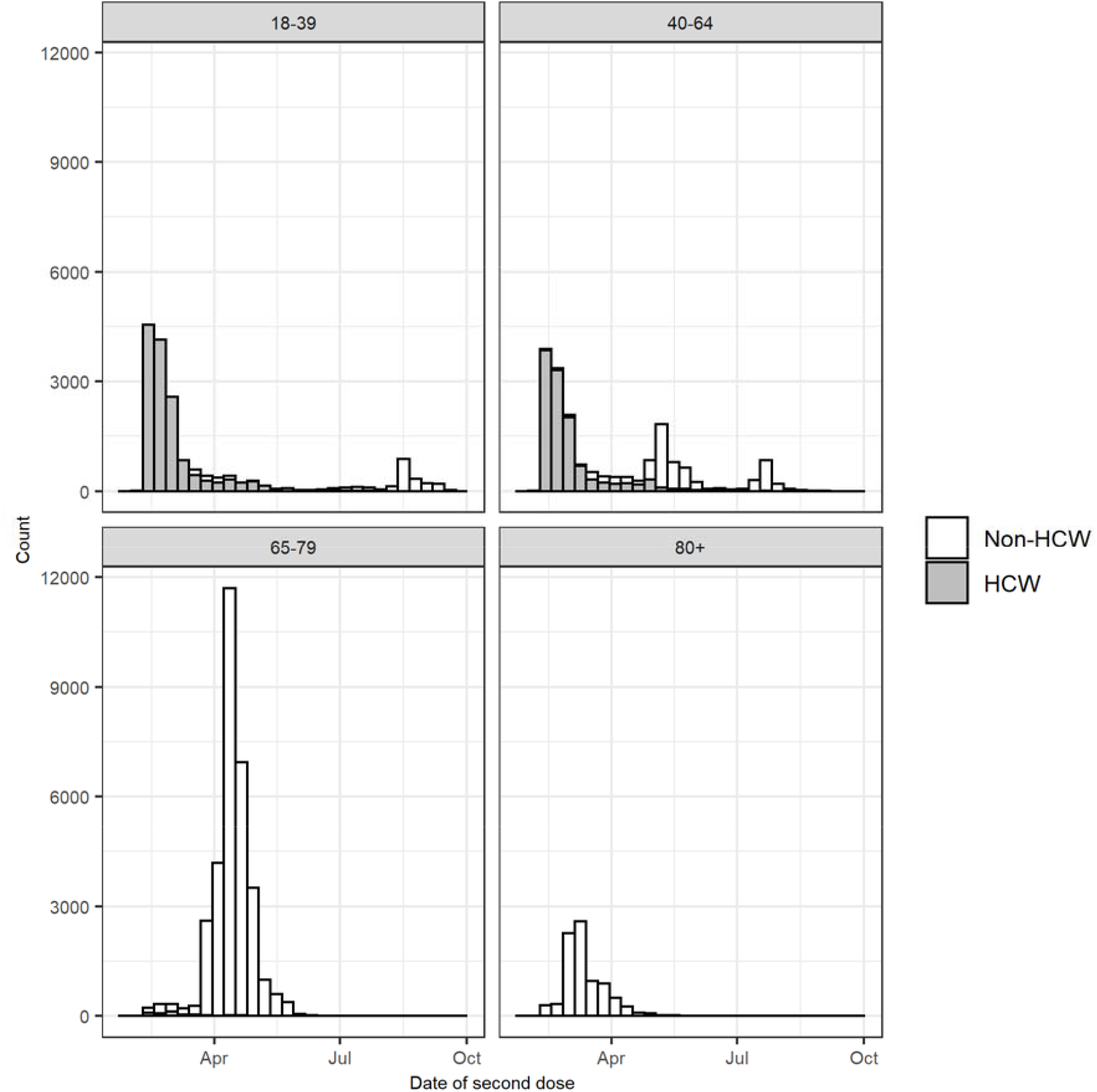
Distribution of second dose timing by age group in matched cases and controls, stacked by healthcare worker status

**Table 1.**
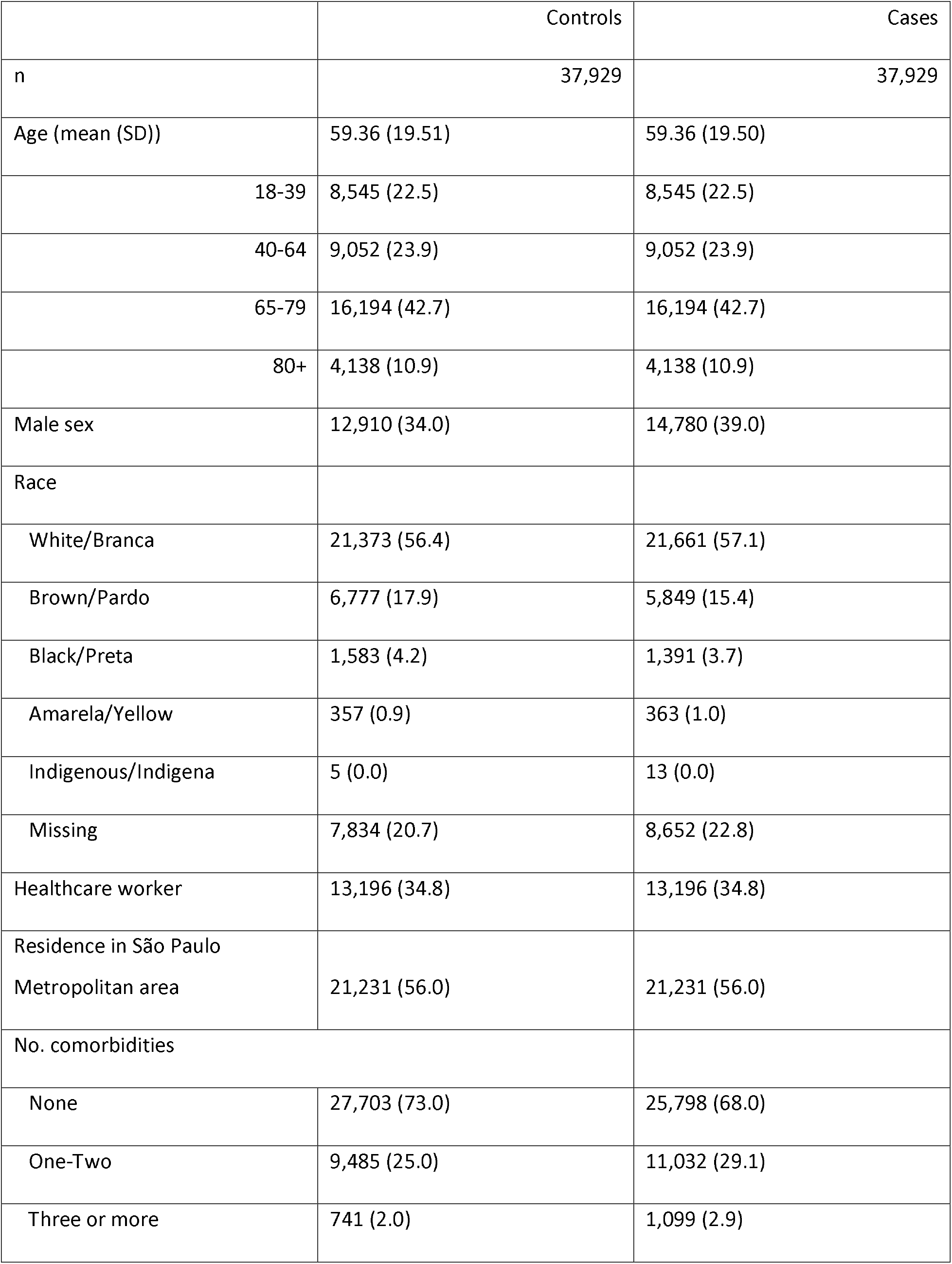
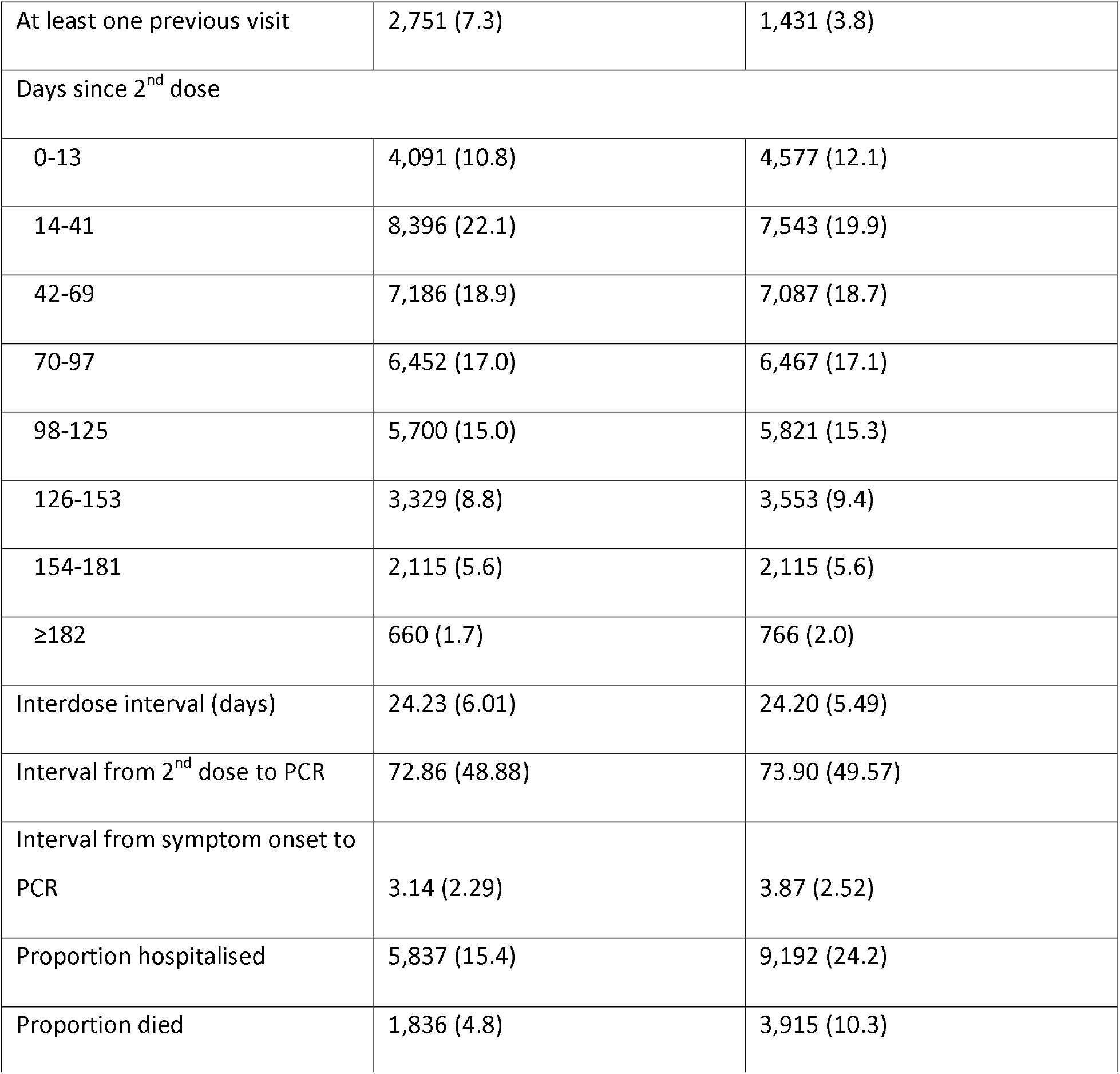
Demographic and clinical characteristics of matched cases and controls

|  | Controls | Cases |
| --- | --- | --- |
| n | 37,929 | 37,929 |
| Age (mean (SD)) | 59.36 (19.51) | 59.36 (19.50) |
| 18-39 | 8,545 (22.5) | 8,545 (22.5) |
| 40-64 | 9,052 (23.9) | 9,052 (23.9) |
| 65-79 | 16,194 (42.7) | 16,194 (42.7) |
| 80+ | 4,138 (10.9) | 4,138 (10.9) |
| Male sex | 12,910 (34.0) | 14,780 (39.0) |
| Race |  |  |
| White/Branca | 21,373 (56.4) | 21,661 (57.1) |
| Brown/Pardo | 6,777 (17.9) | 5,849 (15.4) |
| Black/Preta | 1,583 (4.2) | 1,391 (3.7) |
| Amarela/Yellow | 357 (0.9) | 363 (1.0) |
| Indigenous/Indigena | 5 (0.0) | 13 (0.0) |
| Missing | 7,834 (20.7) | 8,652 (22.8) |
| Healthcare worker | 13,196 (34.8) | 13,196 (34.8) |
| Residence in São Paulo<br>Metropolitan area | 21,231 (56.0) | 21,231 (56.0) |
| No. comorbidities |  |  |
| None | 27,703 (73.0) | 25,798 (68.0) |
| One-Two | 9,485 (25.0) | 11,032 (29.1) |
| Three or more | 741 (2.0) | 1,099 (2.9) |
| At least one previous visit | 2,751 (7.3) | 1,431 (3.8) |
| Days since 2<sup>nd</sup> dose |  |  |
| 0-13 | 4,091 (10.8) | 4,577 (12.1) |
| 14-41 | 8,396 (22.1) | 7,543 (19.9) |
| 42-69 | 7,186 (18.9) | 7,087 (18.7) |
| 70-97 | 6,452 (17.0) | 6,467 (17.1) |
| 98-125 | 5,700 (15.0) | 5,821 (15.3) |
| 126-153 | 3,329 (8.8) | 3,553 (9.4) |
| 154-181 | 2,115 (5.6) | 2,115 (5.6) |
| ≥182 | 660 (1.7) | 766 (2.0) |
| Interdose interval (days) | 24.23 (6.01) | 24.20 (5.49) |
| Interval from 2<sup>nd</sup> dose to PCR | 72.86 (48.88) | 73.90 (49.57) |
| Interval from symptom onset to PCR | 3.14 (2.29) | 3.87 (2.52) |
| Proportion hospitalised | 5,837 (15.4) | 9,192 (24.2) |
| Proportion died | 1,836 (4.8) | 3,915 (10.3) |

### Change in odds of COVID-19 over time since vaccination

The final model selected by AIC for the outcome of symptomatic COVID-19 had a two-way interaction between each of HCW status and age, and time since vaccination. The odds ratio of symptomatic COVID-19 increased with time from vaccination, the pattern of which varied by age group among non-HCWs (Figure 3 and Supplementary Table 3). Among individuals aged ≥80 years, those vaccinated more than 70 days prior to their test date had increased odds of COVID-19 relative to those vaccinated 14-41 days prior (OR 1.43, 95% CI 1.05 to 1.94), and the odds ratio increased over time to 3.32 (95% CI 1.85 to 5.94) after ≥182 days. Overall the odds ratio of COVID-19 associated with time since vaccination was of lower magnitude among 40-64 year olds and 65-79 year-olds, with maximum odds ratios of 1.75 (95% CI 1.04 to 2.96) and 1.48 (95% CI 1.18 to 1.87) in the two age groups respectively. Among 18-39 year-old non-HCWs, significantly increased odds of COVID-19 infection began at 42-69 days (OR 1.45, 95% CI 1.15 to 1.82), increasing to 3.87 (95% CI 2.16 to 6.92) at 154-181 days.

**Figure 3.**
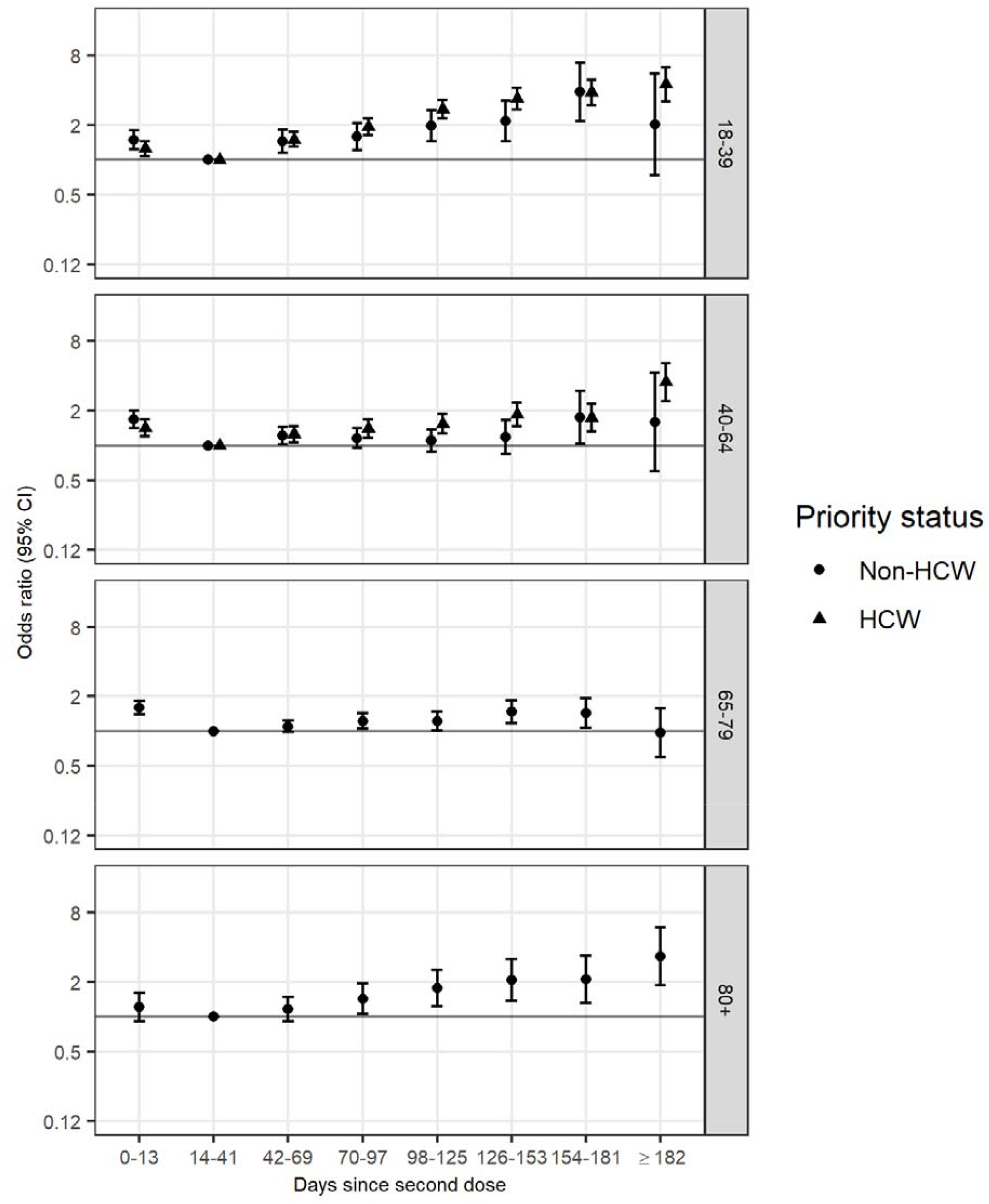
Odds ratio (on a log scale) of symptomatic PCR-confirmed COVID-19 disease against days since vaccination, relative to 14-41 days following vaccination, by age group (rows), for non-healthcare workers (left column) and healthcare workers (right column)

Among HCWs, increased odds of COVID-19 infection began at 42-69 days (among 18-39 year-old HCWs, OR 1.49, 95% CI 1.29 to 1.73; among 40-64 year-old HCWs, OR 1.26, 95% CI 1.07 to 1.48) and the odds ratio increased with time since vaccination (among 18-39 year-old HCWs, OR from 182 days 4.48, 95% CI 3.19 to 6.28; among 40-64 year-old HCWs, OR from 182 days 3.53, 95% CI 2.42 to 5.14). The pattern was similar for 18-39 year-old and 40-64 year-old HCWs, with greater increases among the younger age group. For both age groups, the increase in COVID-19 odds over time was greater among HCWs than non-HCWs (Supplementary Table 3).

In all groups, individuals receiving their second dose between 0 and 13 days before the RT-PCR test had significantly increased odds of COVID-19, indicating the time period before the second dose is maximally effective.

### Change in odds of severe COVID-19 outcomes over time since vaccination

The final model selected by AIC for the outcome of COVID-19 hospitalisation or death included no interactions between HCW status or age and time since vaccination. Longer time from second dose was associated with increased odds of COVID-19 hospitalisation or death, with a significant increase observed starting at 98-125 days (OR 1.40, 95% CI 1.09 to 1.79) (Figure 4 and Supplementary Table 4).

**Figure 4.**
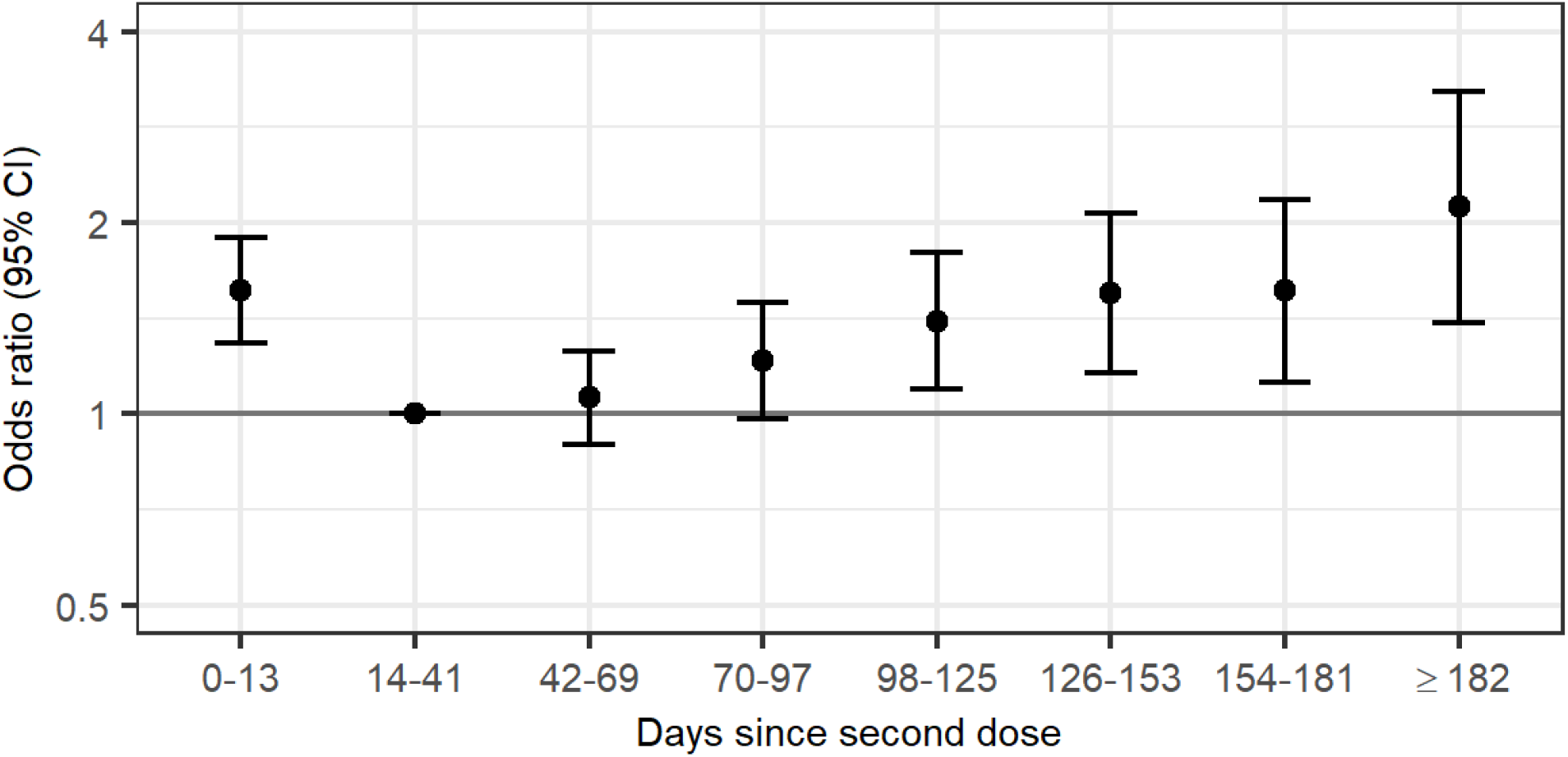
Odds ratio (on a log scale) of PCR-confirmed COVID-19 hospitalisation or death for days since vaccination, relative to 14-41 days following vaccination

### Exploring sources of bias

Estimates of waning in this design can be affected by several sources of bias, which may explain the patterns seen in Figure 3. To examine the potential for such an association, we estimated the association between month of vaccination and COVID-19 among recent vaccinees (Sensitivity Analysis 1). Supplementary Figure 2 shows the odds ratio of COVID-19 by month of vaccination, among individuals receiving an RT-PCR test 14-90 days following their second dose. Among non-HCWs the odds of COVID-19 was significantly lower among individuals vaccinated from May to July compared to April (ORs ranging from 0.47 to 0.76), but those vaccinated in February and March are at otherwise similar risk. On the other hand, early adopters among HCWs appear to be at significantly higher risk of COVID-19 than later adopters (e.g. OR comparing HCWs vaccinated in February to HCWs vaccinated in April, 2.06, 95% CI, 1.58 to 2.67).

Restricting the analysis to HCWs vaccinated in February (Sensitivity Analysis 2) led to a pattern of waning that was later in onset among 40-64 year olds, with significantly increased odds of COVID-19 starting at 182 days post second dose (Supplementary Figure 3). However, the pattern of waning across time among 18-39 year-old HCWs was similar to the primary analysis. Moreover, the pattern of waning was similar for 18-39 year-old HCWs vaccinated in February and those vaccinated in March (Sensitivity Analysis 3). In contrast, 40-64 year-old HCWs vaccinated in March exhibited stronger apparent waning, albeit with low precision. These analyses suggest some robustness for the waning results among 18-39 year-olds, but that bias may be affecting estimates of waning among 40-64 year-old HCWs.

## Discussion

In this study we have shown evidence of an increase in COVID-19 disease and hospitalisation over time since vaccination in individuals receiving two doses of CoronaVac in São Paulo State, Brazil, between February and September 2021. We observed a greater increase in COVID-19 over time since vaccination in younger individuals, and among HCWs compared to non-HCWs. These observed increases in COVID-19 over time can be interpreted as waning of vaccine effectiveness, and such a finding has important implications globally. As of the end of 2021, approximately one quarter of all COVID-19 vaccine doses distributed worldwide have been CoronaVac, the majority of which have been given to individuals in lower- and middle-income countries. Any decrease over time in vaccine effectiveness will direct future vaccination policy and affect future COVID-19 mortality.

The difference in magnitude of apparent waning by age (Figure 3), with largest odds ratios observed in the youngest age group, has several potential explanations. Immune response to vaccination is lower in older individuals(20,21), particularly those above 80 years old, which could lead to earlier waning when a protective threshold is crossed.

Differences in the apparent magnitude of waning by age could be explained by differences in initial effectiveness: for example, a drop in VE from 90% to 80% translates to an OR of 2, whereas a drop in VE from 50% to 40% translates to an OR of 1.2. In particular, this phenomenon could explain the large magnitude of waning against COVID-19 among younger individuals for whom initial vaccine effectiveness was likely higher(14).

We explored various sources of bias that could explain the patterns of observed waning. HCWs vaccinated earliest may be at highest risk for SARS-CoV-2 infection, due to occupational exposure. Since HCWs were vaccinated earliest, they are over-represented in later time periods since vaccination, potentially leading to apparent waning. We found suggestive evidence of an association between time of vaccine uptake and COVID-19 risk, meaning that in particular HCWs vaccinated earlier appeared to be at higher COVID-19 risk. Consequently, after adjusting for calendar time of the RT-PCR test, it is impossible to distinguish vaccination time from time since vaccination. We found that this association to be less apparent among non-HCWs, suggesting that these estimates are less affected by such bias. This issue represents a major methodological challenge for observational studies of vaccine waning, and the bias will manifest itself in different ways depending on the dynamics of the epidemic, the correlation between vaccine uptake and COVID-19 risk, and the way in which vaccine roll-out was conducted.

Another potential explanation for apparent waning is depletion of susceptibles (i.e. infection of HCWs prior to their vaccination between February and April) that could result in the vaccine appearing to provide increased protection among later vaccinees who had additional immune response to prior infection. The extent of waning was reduced when restricting to those vaccinated in February among 40-64 year olds but not 18-39 year olds, suggesting that depletion of susceptibles does not explain the waning observed in the latter group. When restricting to individuals vaccinated in March, apparent waning increased among 40-64 year olds, albeit with low precision. These differing results, which should be similar if the increased odds of infection were the result of waning alone, suggest that bias affected our estimates, but did not account for the observed waning overall.

This study is one of a growing number of observational studies assessing waning of vaccine effectiveness, all of which are potentially affected by the same sources of bias. Studies that estimate vaccine effectiveness by comparing vaccinated individuals to unvaccinated individuals at the same point in time(5,7,9) (ref Cohn et al, Tartof et al, Chemaitelly et al) can be affected by depletion of susceptibles bias, leading to overestimation of waning. In addition, none of the current study designs appear to account for any difference in risk by calendar time of vaccination; nonetheless, Mizrahi et al(4) found increased incidence rate of breakthrough infections among early vaccinees compared to late vaccinees, concluding that this provided evidence of waning. A similar case-control study conducted in Israel(8) among individuals who received two doses of BNT162b2 found a 2.37- to 2.82-fold increase in odds of COVID-19 for 90 to ≥180 days following series completion, compared to days 21-90. While our estimates cannot be directly compared due to the different vaccines and contexts, we found up to 4-fold increase in COVID-19 odds 182 days after vaccination, and waning that differed by age and HCW status.

A major strength of this study is our ability to distinguish those vaccinated as healthcare workers against those vaccinated under some other priority group. Given the strong associations between HCW status, age, vaccine timing, and SARS-CoV-2 risk, studies of waning effectiveness must account for the presence of HCWs. Previous studies have done so by restricting to HCWs(6), adjusting for HCW status(7), or using frequency of prior PCR testing as a proxy for occupational exposure (REFs), while other observational studies of vaccine effectiveness have excluded HCWs(22,23). However, the authors are not aware of evidence for difference in apparent waning by HCW status. Current and future studies of vaccine effectiveness must be designed to address bias by HCWs and other high-risk individuals who are prioritised for vaccination, and preferably to examine patterns within risk groups.

There are several limitations of this study. We did not have data on the variant of concern for each case included in the study. Therefore, some of the waning observed may be due to decreased effectiveness of CoronaVac against Delta compared to Gamma. This design is less affected by this issue than a design that estimates vaccine effectiveness over calendar time, but nonetheless, later calendar times are over-represented in later times since vaccination. By design, we did not estimate vaccine effectiveness, which would require a comparison with unvaccinated individuals, but rather change in odds of disease post-vaccination relative to a period shortly after the second dose. Therefore, we are unable to provide estimates of vaccine effectiveness after six months. In our analysis of hospitalisation or death, changing hospitalisation triage criteria over time and different thresholds for hospitalisation among vaccinated patients may have contributed to our estimates of waning. Our sensitivity analyses suggested bias but were unable to fully correct for it. For example, we offer an alternative explanation for observed early waning in HCWs, namely that early adopters were at higher COVID-19 risk. However, waning of humoral immune response has been observed on this timescale (ref Saure), so the extent to which increased SARS-CoV-2 infection risk among early vaccines contributes to apparent waning remains unclear. Finally, even within non-HCWs, CoronaVac was only available to certain high-risk groups before May, leading to over-representation of higher risk individuals and increased COVID-19 odds in later time periods.

Our results support the public health decision made by the WHO-SAGE group on recommending a third dose in the elderly who received two doses of CoronaVac (REF SAGE). This was expected based on the decreased effectiveness observed for two-dose schedules in this age group for inactivated vaccines [ref]. Additionally, we observed a waning of protection among those <60 years, mainly represented by HCWs. The recommendation for booster doses for all adults still lacks robust evidence, and our analyses based on HCWs support this decision. Some countries with high coverage of two doses, such as Brazil and Chile, had already recommended a booster dose for all adults. Additional analyses are needed for the ideal timing of booster doses, whether homologous and heterologous schedules, and the role of new variants in this context.

We have provided evidence that moderate and severe covid-19 outcomes increased over time following CoronaVac primary series completion, and have suggested several sensitivity analyses that could be conducted to understand bias in such observational studies.

## Data Availability

Deidentified databases as well as the R codes will be deposited in the repository https://github.com/juliocroda/VebraCOVID-19

## Author contributions

All authors conceived the study. MDTH completed analyses with guidance from OTR, NED, JRA, DATC, AIK, and JC. MSST, OFPP, OTR and MDTH curated and validated the data. MDTH wrote the first draft of the manuscript. TLD, RCP, OFPP, EFMV, MA, RS, JCG, WNA provided supervision. MLL and MD edited and reviewed the manuscript. All authors contributed to, and approved, the final manuscript. JC is the guarantor. The corresponding author attests that all listed authors meet authorship criteria and that no others meeting the criteria have been omitted.

## Ethics approval

The study was approved by the Ethical Committee for Research of Federal University of Mato Grosso do Sul (CAAE: 43289221.5.0000.0021).

## Public and Patient Involvement

Members of the public or patients were not involved in setting the research question or the outcome measures, nor were they involved in developing plans for the design of the study. No patients were asked to advise on interpretation or writing up of results.

## Transparency statement

The lead author affirms that the manuscript is an honest, accurate, and transparent account of the study being reported; that no important aspects of the study have been omitted; and that any discrepancies from the study as originally planned have been explained.

## Dissemination declaration

Results will be disseminated to the public in São Paulo and across Brazil. It is not possible to disseminate results to individuals who were selected into the study due to anonymisation of the data.

## Acknowledgements

We are grateful for the Pan American Health Organization’s support and the São Paulo State in making the databases available for analysis. JC is supported by the Oswaldo Cruz Foundation (Edital Covid-19 – resposta rápida: 48111668950485). OTR is funded by a Sara Borrell fellowship (CD19/00110) from the Instituto de Salud Carlos III. OTR acknowledges support from the Spanish Ministry of Science and Innovation through the Centro de Excelencia Severo Ochoa 2019-2023 Program and from the Generalitat de Catalunya through the CERCA Program. This work was supported by grant R01-AI139761 from the National Institute for Allergy and Infectious Diseases to N.E.D.

