## Supplementary Material for "Change in COVID-19 risk over time following vaccination with CoronaVac: A test-negative case-control study"

p2: Supplementary Table 1

p4: Supplementary Figure 1

p5: Supplementary Table 2

p6: Supplementary Table 3

p8: Supplementary Table 4

p9: Supplementary Figure 2

p10: Supplementary Figure 3

|  | Item No | Recommendation | Page No |
| --- | --- | --- | --- |
| **Title and abstract** | 1 | (*a*) Indicate the study’s design with a commonly used term in the title or the abstract | 1 |
|  |  | (*b*) Provide in the abstract an informative and balanced summary of what was done and what was found | 2 |
| Introduction | | | |
| Background/rationale | 2 | Explain the scientific background and rationale for the investigation being reported | 4 |
| Objectives | 3 | State specific objectives, including any prespecified hypotheses | 4 |
| Methods | | | |
| Study design | 4 | Present key elements of study design early in the paper | 5 |
| Setting | 5 | Describe the setting, locations, and relevant dates, including periods of recruitment, exposure, follow-up, and data collection | 4-5 |
| Participants | 6 | (*a*) Give the eligibility criteria, and the sources and methods of case ascertainment and control selection. Give the rationale for the choice of cases and controls | 5 |
|  |  | (*b*) For matched studies, give matching criteria and the number of controls per case | 5 |
| Variables | 7 | Clearly define all outcomes, exposures, predictors, potential confounders, and effect modifiers. Give diagnostic criteria, if applicable | 5-6 |
| Data sources/ measurement | 8* | For each variable of interest, give sources of data and details of methods of assessment (measurement). Describe comparability of assessment methods if there is more than one group | 4 |
| Bias | 9 | Describe any efforts to address potential sources of bias | 5-7 |
| Study size | 10 | Explain how the study size was arrived at | 8; Fig 1 |
| Quantitative variables | 11 | Explain how quantitative variables were handled in the analyses. If applicable, describe which groupings were chosen and why | 5-6 |
| Statistical methods | 12 | (*a*) Describe all statistical methods, including those used to control for confounding | 5-6 |
|  |  | (*b*) Describe any methods used to examine subgroups and interactions | 5-6 |
|  |  | (*c*) Explain how missing data were addressed | 5-6 |
|  |  | (*d*) If applicable, explain how matching of cases and controls was addressed | 6 |
|  |  | (*e*) Describe any sensitivity analyses | 6-7 |
| Results | | | |
| Participants | 13* | (a) Report numbers of individuals at each stage of study—eg numbers potentially eligible, examined for eligibility, confirmed eligible, included in the study, completing follow-up, and analysed | 8; Fig 1, Supp Mat |
|  |  | (b) Give reasons for non-participation at each stage | Fig 1 |
|  |  | (c) Consider use of a flow diagram | Fig 1 |
| Descriptive data | 14* | (a) Give characteristics of study participants (eg demographic, clinical, social) and information on exposures and potential confounders | 8; Table 1 |
|  |  | (b) Indicate number of participants with missing data for each variable of interest | Table 1 |
| Outcome data | 15* | Report numbers in each exposure category, or summary measures of exposure | Table 1; Supp Mat |

Supplementary Table 1. STROBE checklist

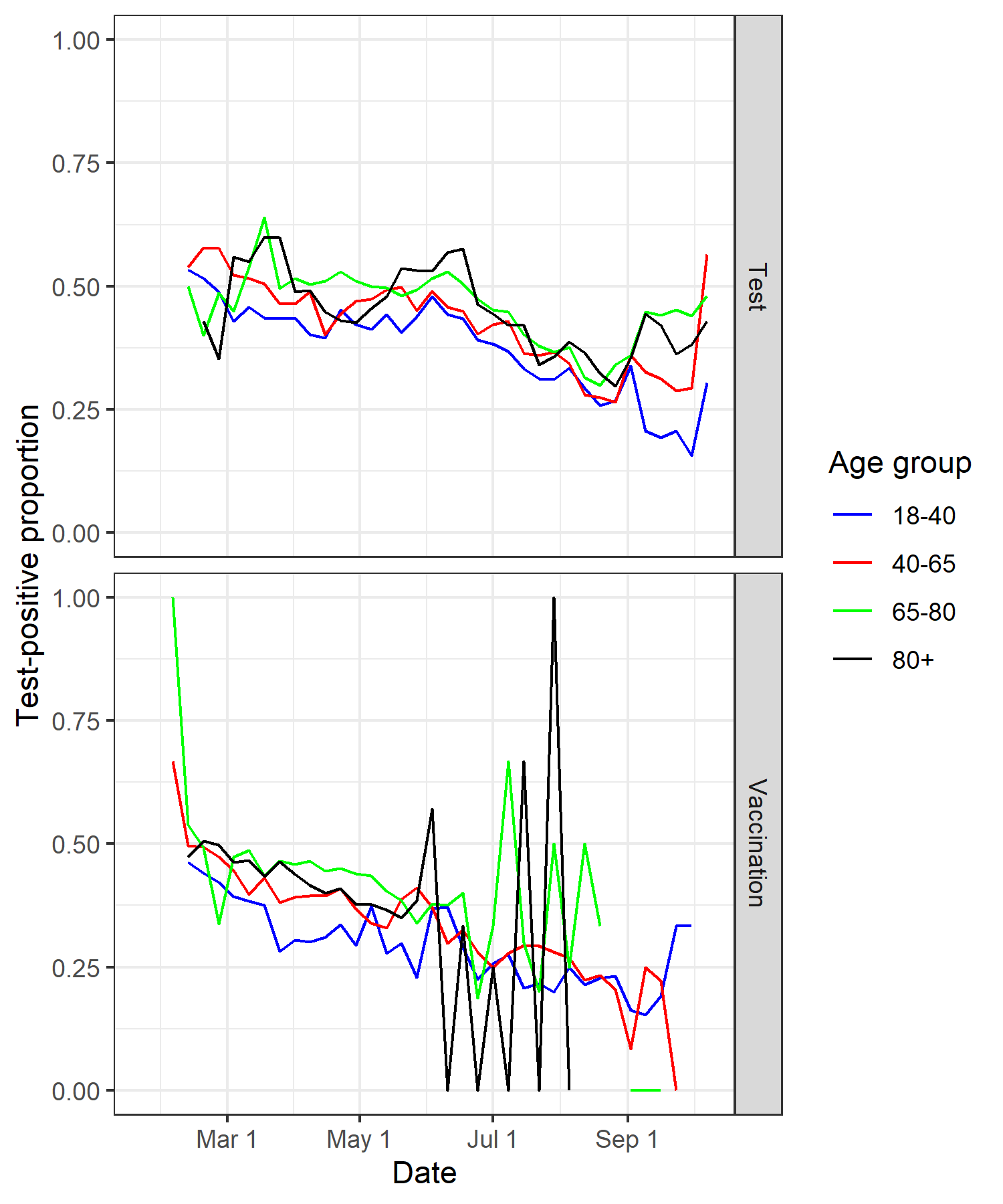

Supplementary Figure 1. Proportion of RT-PCR tests that were positive by week of test performance (top) and by week of primary series completion (bottom), by age group, in the study population.

|  | Case  0-14 | Case  14-41 | Case  42-69 | Case  70-97 | Case  98-125 | Case 126-153 | Case 154-181 | Case ≥182 |
| --- | --- | --- | --- | --- | --- | --- | --- | --- |
| Control 0-14 | 2764 | 961 | 149 | 134 | 44 | 24 | 11 | 4 |
| Control 14-41 | 1392 | 5149 | 1258 | 341 | 190 | 51 | 9 | 6 |
| Control 42-69 | 211 | 1016 | 4485 | 1048 | 315 | 82 | 22 | 7 |
| Control 70-97 | 164 | 240 | 870 | 3826 | 1077 | 186 | 77 | 12 |
| Control 98-125 | 33 | 148 | 231 | 925 | 3457 | 699 | 171 | 36 |
| Control 126-153 | 9 | 19 | 69 | 149 | 581 | 2002 | 439 | 61 |
| Control 154-181 | 4 | 8 | 21 | 35 | 137 | 450 | 1203 | 257 |
| Control ≥182 | 0 | 2 | 4 | 9 | 20 | 59 | 183 | 383 |

Supplementary Table 2. Number of pairs by case and control days since receipt of second dose (primary analysis)

|  |  | Non-HCWs | HCWs |
| --- | --- | --- | --- |
| Variable | Age subgroup | Odds ratio (95% CI) | |
| Age | All | 1.00 (0.99-1.01) | |
| Race |  |  | |
| White/Branca |  | Ref | |
| Brown/Pardo |  | 0.85 (0.81-0.88) | |
| Black/Preta |  | 0.84 (0.78-0.91) | |
| Yellow/Amarela |  | 0.98 (0.84-1.14) | |
| Indigenous |  | N/A | |
| Missing |  | 1.09 (1.05-1.13) | |
| Any prior ARI |  | 0.47 (0.44-0.50) | |
| Female sex |  | 0.81 (0.78-0.83) | |
| Number comorbidities |  |  | |
| None |  | Ref | |
| One-Two |  | 1.36 (1.31-1.41) | |
| Three or more |  | 1.78 (1.61-1.96) | |
| Days since second dose (relative to 14-41) |  |  |  |
| 0-14 | 18-39 | 1.48 (1.23-1.78) | 1.25 (1.08-1.45) |
| 42-69 |  | 1.45 (1.15-1.82) | 1.49 (1.29-1.73) |
| 70-97 |  | 1.59 (1.21-2.09) | 1.93 (1.64-2.27) |
| 98-125 |  | 1.97 (1.44-2.68) | 2.74 (2.30-3.28) |
| 126-153 |  | 2.17 (1.44-3.26) | 3.39 (2.73-4.21) |
| 154-181 |  | 3.87 (2.16-6.92) | 3.83 (2.97-4.94) |
| ≥182 |  | 2.03 (0.73-5.58) | 4.48 (3.19-6.28) |
| 0-14 | 40-64 | 1.68 (1.42-2.00) | 1.43 (1.20-1.69) |
| 42-69 |  | 1.22 (1.02-1.46) | 1.26 (1.07-1.48) |
| 70-97 |  | 1.16 (0.95-1.41) | 1.40 (1.17-1.68) |
| 98-125 |  | 1.10 (0.88-1.38) | 1.54 (1.26-1.88) |
| 126-153 |  | 1.18 (0.85-1.65) | 1.85 (1.46-2.35) |
| 154-181 |  | 1.75 (1.04-2.96) | 1.74 (1.32-2.28) |
| ≥182 |  | 1.59 (0.60-4.24) | 3.53 (2.42-5.14) |
| 0-14 | 65-79 | 1.60 (1.41-1.82) | - |
| 42-69 |  | 1.10 (0.99-1.23) | - |
| 70-97 |  | 1.23 (1.05-1.43) | - |
| 98-125 |  | 1.23 (1.02-1.48) | - |
| 126-153 |  | 1.48 (1.18-1.87) | - |
| 154-181 |  | 1.44 (1.07-1.93) | - |
| ≥182 |  | 0.97 (0.60-1.57) | - |
| 0-14 | 80+ | 1.22 (0.92-1.61) | - |
| 42-69 |  | 1.16 (0.91-1.48) | - |
| 70-97 |  | 1.43 (1.05-1.94) | - |
| 98-125 |  | 1.77 (1.23-2.54) | - |
| 126-153 |  | 2.07 (1.36-3.16) | - |
| 154-181 |  | 2.10 (1.32-3.37) | - |
| ≥182 |  | 3.32 (1.85-5.94) | - |

Supplementary Table 3. Adjusted odds ratios for COVID-19

| Variable | Odds ratio (95% CI) |
| --- | --- |
| Age | 1.03 (1.00-1.05) |
| Race |  |
| White/Branca | Ref |
| Brown/Pardo | 0.70 (0.64-0.77) |
| Black/Preta | 0.77 (0.65-0.91) |
| Yellow/Amarela | 1.22 (0.93-1.60) |
| Indigenous | N/A |
| Missing | 0.67 (0.61-0.74) |
| Any prior ARI | 0.29 (0.22-0.37) |
| Female sex | 0.63 (0.59-0.67) |
| Number comorbidities |  |
| None | Ref |
| One-Two | 3.13 (2.91-3.36) |
| Three or more | 5.51 (4.78-6.35) |
| Days since second dose (relative to 14-41) |  |
| 0-14 | 1.56 (1.29-1.89) |
| 42-69 | 1.06 (0.90-1.25) |
| 70-97 | 1.21 (0.98-1.50) |
| 98-125 | 1.40 (1.09-1.79) |
| 126-153 | 1.55 (1.16-2.07) |
| 154-181 | 1.56 (1.12-2.18) |
| >=182 | 2.12 (1.39-3.22) |

Supplementary Table 4. Adjusted odds ratios for COVID-19 hospitalisation or death

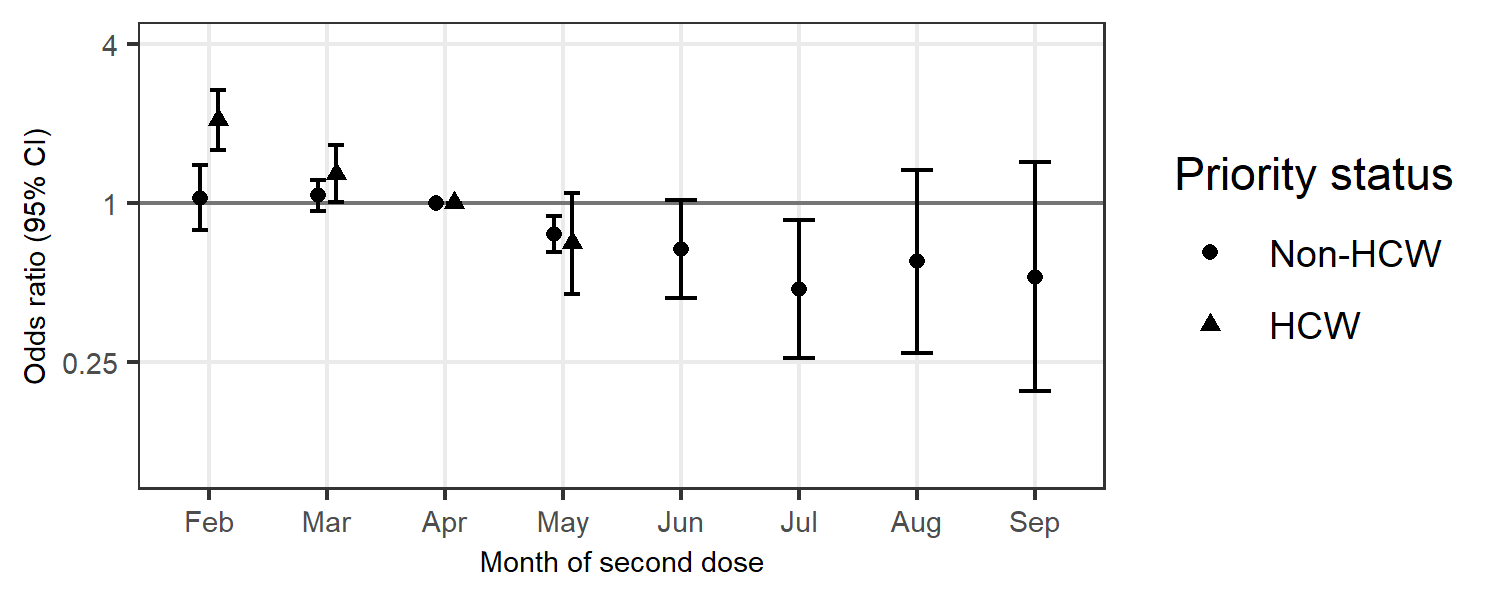
Supplementary Figure 2. Odds ratio (on a log scale) of symptomatic COVID-19 by month of second dose, among individuals receiving an RT-PCR test within 14-90 days of their second dose, for HCWs (top row) and non-HCWs (bottom row)

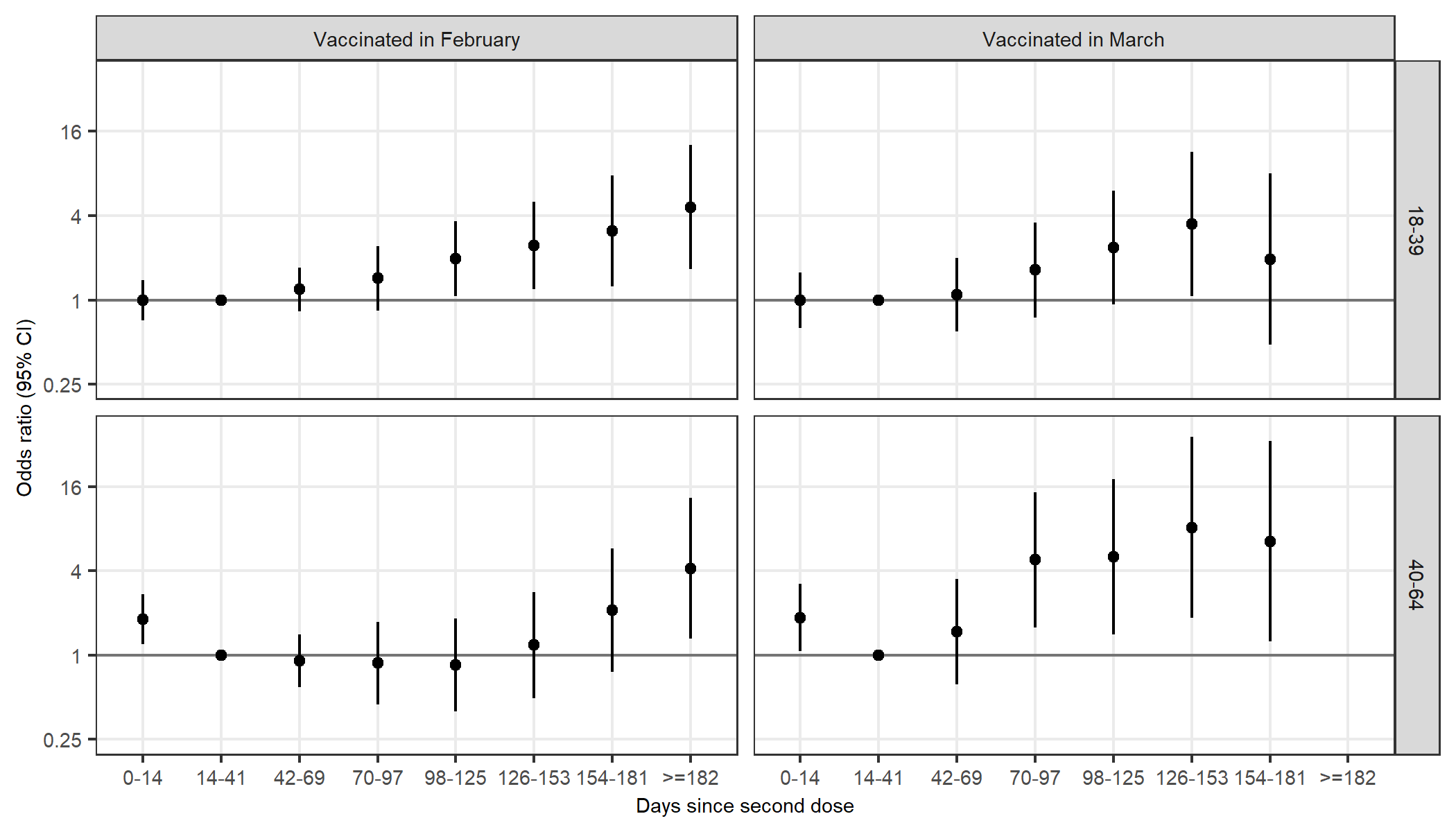
Supplementary Figure 3. Odds ratio (on a log scale) of symptomatic COVID-19 for days since vaccination, relative to 14-41 days following vaccination for HCWs who received their second dose in February 2021 (left column) and March 2021 (right column), by age group (rows).
